# Understanding unexpected results from randomized clini□cal trials Does coffee reduce atrial fibrillation recurrences?

**DOI:** 10.64898/2026.04.13.26350787

**Authors:** James M Brophy

## Abstract

**Objective:** To explore the interpretation of *unexpected* results from a randomized controlled trial (RCT).

**Study Design and Setting:** Adjunctive frequentist (power and type□M error) and Bayesian analyses were performed on a recently published RCT reporting a statistically significant relative risk reduction (p <0.01) for caffeinated coffee drinkers compared with abstinence on atrial fibrillation (AF) recurrence. Individual patient data for the Bayesian survival models were reconstructed from the RCT published material and priors informed by the RCT power calculations.

**Results:** The original RCT design had limited power for realistic effect sizes, increasing susceptibility to type□M (magnitude) error. Bayesian analyses also tempered the benefit for caffeinated coffee implied by standard statistical analysis resulting in only modest probabilities of clinically meaningful risk reductions (e.g., hazard ratio < 0.9 of 88% or a risk difference > 2% of 82%).

**Conclusions:** Supplemental frequentist and Bayesian approaches can provide robustness checks for *unexpected* RCT findings, providing contextualization, clarifying distinctions between statistical and clinical significance, and guiding replication needs.

**Highlights:**

- Randomized controlled trial (RCT) results may be *unexpected* and challenge prior beliefs
- Supplemental frequentist and Bayesian analyses can clarify interpretation of surprising findings
- Power and type□M error assessments help evaluate design adequacy for realistic effects
- Bayesian posterior probabilities provide additional nuanced insights into contextulaization and clinical significance

## Introduction

Results from well performed, unbiased randomized controlled trials (RCT) are generally assumed to provide ground truth and consequently there has been little discussion about how to interpret “unexpected” or “surprising” results. As an example, a recent RCT, “Caffeinated Coffee Consumption or Abstinence to Reduce Atrial Fibrillation The DECAF Randomized Clinical Trial”(1) reported that following successful cardioversion for atrial fibrillation (AF), allocation to consumption of caffeinated coffee averaging 1 cup a day was associated with less recurrence of AF or atrial flutter compared with abstinence from coffee and caffeinated products. Historically caffeine has been seen as proarrhythmic and therefore this result is likely surprising to most physicians. The authors explicitly acknowledge this historical reality stating “Caffeinated coffee has traditionally been considered proarrhythmic”(1) but claim that coffee’s role in atrial fibrillation is actually uncertain.

Supporting this opinion, The DECAF authors(1) refer to a previous randomized crossover trial(2) that was not of atrial fibrillation patients but rather of 100 ambulatory patients in which the caffeinated group did not show an increase in atrial ectopy (rate ratio, 1.09; 95% confidence interval [CI], 0.98 to 1.20; P=0.10). However, given this confidence interval, one should be careful to not confuse absence of evidence for evidence of absence. Also uncertain is whether caffeine’s effect on atrial ectopy is a suitable surrogate for its effect on atrial fibrillation. The DECAF authors(1) also cited a meta-analysis of 12 observational studies(3) that reported no conclusive evidence for or against an association between caffeine intake and incident atrial fibrillation (OR 0.95 (0.84–1.06) but cautiously note “observational studies are prone to confounding,and whether these findings are biased by systematic differences between coffee and noncoffee drinkers is unclear”.

Given this lack of conclusive evidence, DECAF(1) was designed to investigate the effect of caffeinated coffee consumption versus abstinence on atrial fibrillation recurrence following successful cardioversion.

## Methods

### DECAF Trial Design

DECAF(1) was a randomized controlled trial of 200 regular coffee drinkers with a recent successful cardioversion for atrial fibrillation (AF). Patients were randomized to either continue their usual caffeinated coffee consumption (median 1 cup/day) or to abstain from all caffeine products (decaf group). The primary outcome was recurrence of atrial fibrillation or atrial flutter within 6 months. The sample size calculations were based on assuming “… a 50% incidence of AF recurrence within 6 months following cardioversion. A clinically relevant effect size was assumed to approximate the effectiveness of commonly prescribed antiarrhythmic drugs for recurrent AF after cardioversion. To provide 80% power to detect a minimum 41% reduced relative hazard of AF, we enrolled 200 patients (**100 per group**) assuming a 1:1 randomization scheme, a potential 10% loss to follow-up, and .05 2-tailed α level.”(1)

### Statistical methods

To complement the published statistical methods, adjunctive frequentist and Bayesian analyses have been performed. Frequentist M error analyses were performed using the retrodesign package(4). For Bayesian analyses, treatment effect and baseline risk priors were specified as normal distributions on the log-odds scale with model parameters estimated from the reported power calculations(1). The baseline control prior was mean centered at the DECAF(1) belief of a 50% recurrence risk (0 on the logit scale) with a standard deviation of 1.5, indicating a weakly informative prior. Similarly, the prior for the treatment effect was centered at a 41% reduction (29.5% or -0.871 on the logit scale) with an assumed standard deviation of 0.5, reflecting slightly higher confidence in our ability to estimate effect size variability. The reasonableness of these priors was assessed through prior predictive checks which demonstrated that these priors adequately reflect the exisitng uncertainty with a wide range of possible effect sizes, principally with an expected benefit (93.5% probability), but a small probability of harm (6.5%), for the intervention versus control.

In addition to priors, Bayesian survival analysis requires the individual patient data (IPD). Cumulative incidence data were available in graphical format in the original publication(1) and were extracted with WebPlotDigitizer(5). The data were then transformed into survival format to generate a Kaplan–Meier (KM) plot using the Guyot(6) algorithm that incorporates numbers at risk and event counts. This was implemented using the IPDfromK package(7) which provides a robust and validated approach for reconstructing IPD from published cumulative incidence curves for secondary analyses.

Bayesian models with binomial likelihood and a logit link function, appropriate for binary outcome data, were analyzed with the brms package(8). This is a high-level front-end interface for Stan(9), a probabilistic programming language that employs Hamiltonian Monte Carlo (HMC) sampling with the No-U-Turn Sampler (NUTS) algorithm that permits efficient exploration of posterior distributions. As mentionned previously, priors were based on the DECAF(1) power calculation. Posterior estimation was performed via the cmdstanr package(10), which interfaces directly with Stan and provides enhanced computational and diagnostic capabilities. Four chains were run with 2,000 warm-up and 6,000 sampling iterations per chain, using adapt_delta = 0.999 and max_treedepth = 15 to eliminate divergences. Convergence was evaluated using the Gelman–Rubin statistic 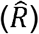 and effective sample size diagnostics. Posterior summaries are reported as medians with 95% credible intervals (CrI). Prior and posterior distributions were visualized using the ggplot2(11) and ggdist(12) packages.

Analyses were performed within the R(13) ecosystem, and the statistical code is available online (https://github.com/brophyj/decaf.git).

## Results

### Critique of DECAF Design

The published DECAF(1) methods are problematic for several reasons.

1. Since the methods section precedes results, stating that 1:1 randomization of 200 subjects will produce two equal groups is problematic. Based on binomial probability theory, there is only a 5.7% chance of achieving this perfectly balanced split (Figure 1).
2. Given the prevailing prior uncertainty, designing a trial to detect a very large risk reduction is overly optimistic. The calculated 80% power estimate assumed a 41% reduction applied to a 50% baseline rates, meaning the intervention is expected to reduce recurrence by an absolute 20.5%. Few cardiology interventions produce such dramatic effects. What would be the power with a sample size of 200 to detect a more modest and realistic risk reduction, say a 15% relative reduction (i.e. from 50% to 42.5%)? The answer is 24% (Figure 2, left panel), illustrating the study was severely underpowered to detect smaller more realistic effect sizes and therefore any statistically significant result is likely to be an exaggeration of the true effect, a Type M (magnitude) error(14). Thus the observed −17% risk difference may be overstating the true benefit, possibly by a factor of two (Figure 2, right panel). The published result must therefore be interpreted cautiously. Further exploration of the probability for a clinically meaningful result awaits a Bayesian analysis.
3. DECAF didn’t specify which arm (abstinence or caffeinated) is hypothesized to reduce the risk. This ambiguity creates interpretive flexibility as whichever arm shows benefit could be claimed as success. Such directional absence hinders inference and inflates the risk of misleading conclusions. This lack of statistical clarity is evident not only in the published article but also in the trial protocol and trial registration documentation at ClinicalTrials.gov.

**Figure 1.**
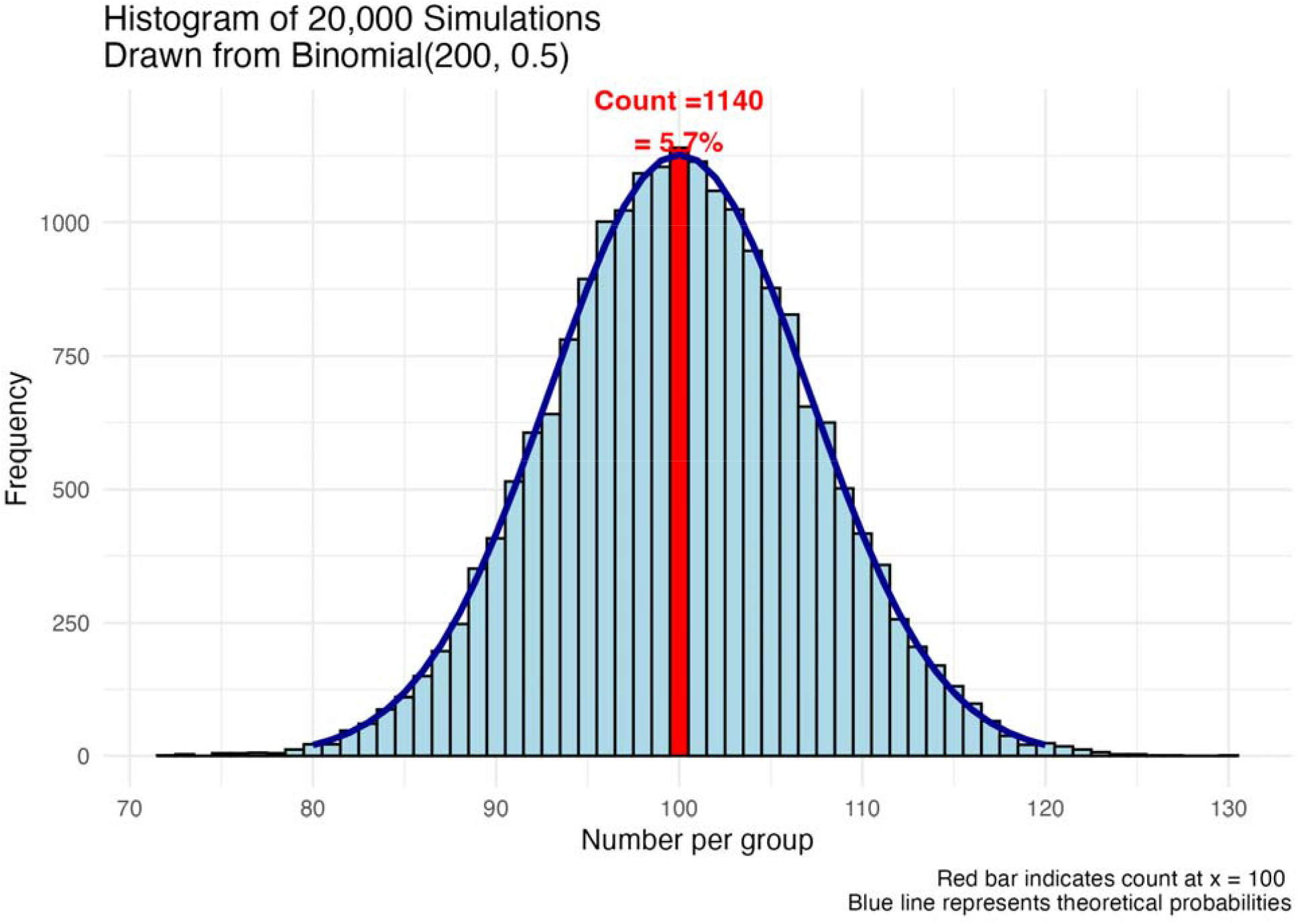
Distribution of group sizes for n=200 randomized 1:1 into two groups. The probability of a perfectly balanced 100/100 split is only 5.7%.

**Figure 2.**
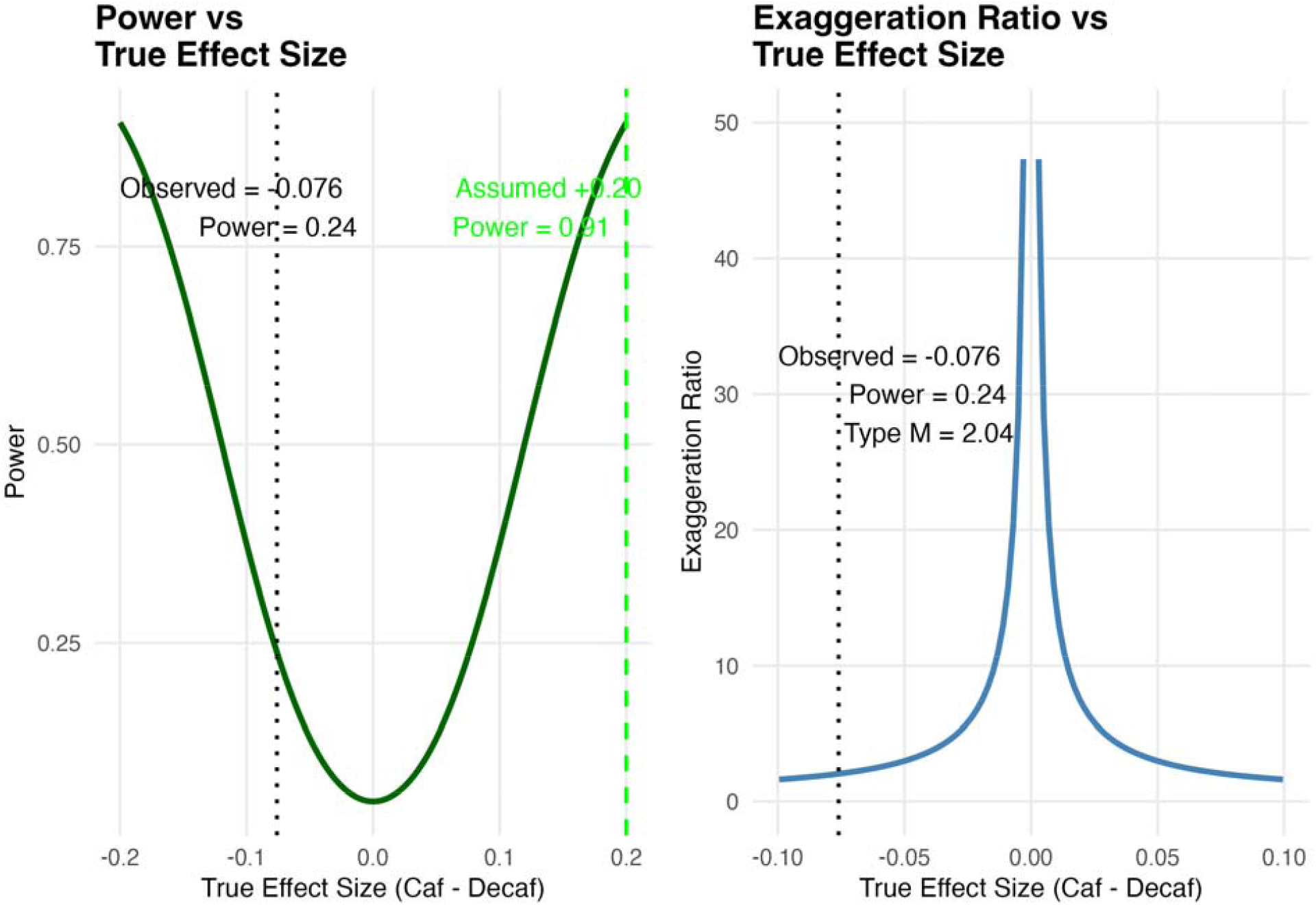
Left panel - Power curves for DECAF trial (n=200) to detect various relative risk reductions from a 50% baseline recurrence rate. The design has only 24% power to detect a 15% relative risk reduction(RRR). Right panel - If the true effect is a 15% RRR, it is likely that any statistically significant result from this underpowered study will overstate the true effect by about two-fold.

### Bayesian relative risk reduction

In Bayesian reanalyses of completed studies, prior beliefs can sometimes be derived from the power calculations. For example, the DECAF(1) authors assumed a baseline recurrence risk of 50% with a 41% relative risk reduction and therefore a 29.5% risk in the intervention group (0.50 - 0.50*0.41 = 0.295). However we are still faced with the previous directional identification problem in the projected improvement. Did DECAF(1) authors believe that the risk reduction would be with abstinence or caffeinated exposure? Given the prevailing historical belief of caffeine as a proarrhythmic agent and given that the study population was comprised initially of coffee drinkers, with the intervention being abstinence we have assumed the proposed relative risk reduction was in favor of the decaffeinated group.

The success of the IPD extraction, required for the Bayesian survival analysis, is evident from a comparison between the published and reconstructed cumulative incidence plots (Figure 3). The IPD calculated frequentist survival analysis results, HR 0.62 (95% CI 0.43–0.91), closely matches the published results, HR 0.61 (95% CI 0.42-0.89)(1), confirming the accuracy of the IPD reconstruction process.

**Figure 3:**
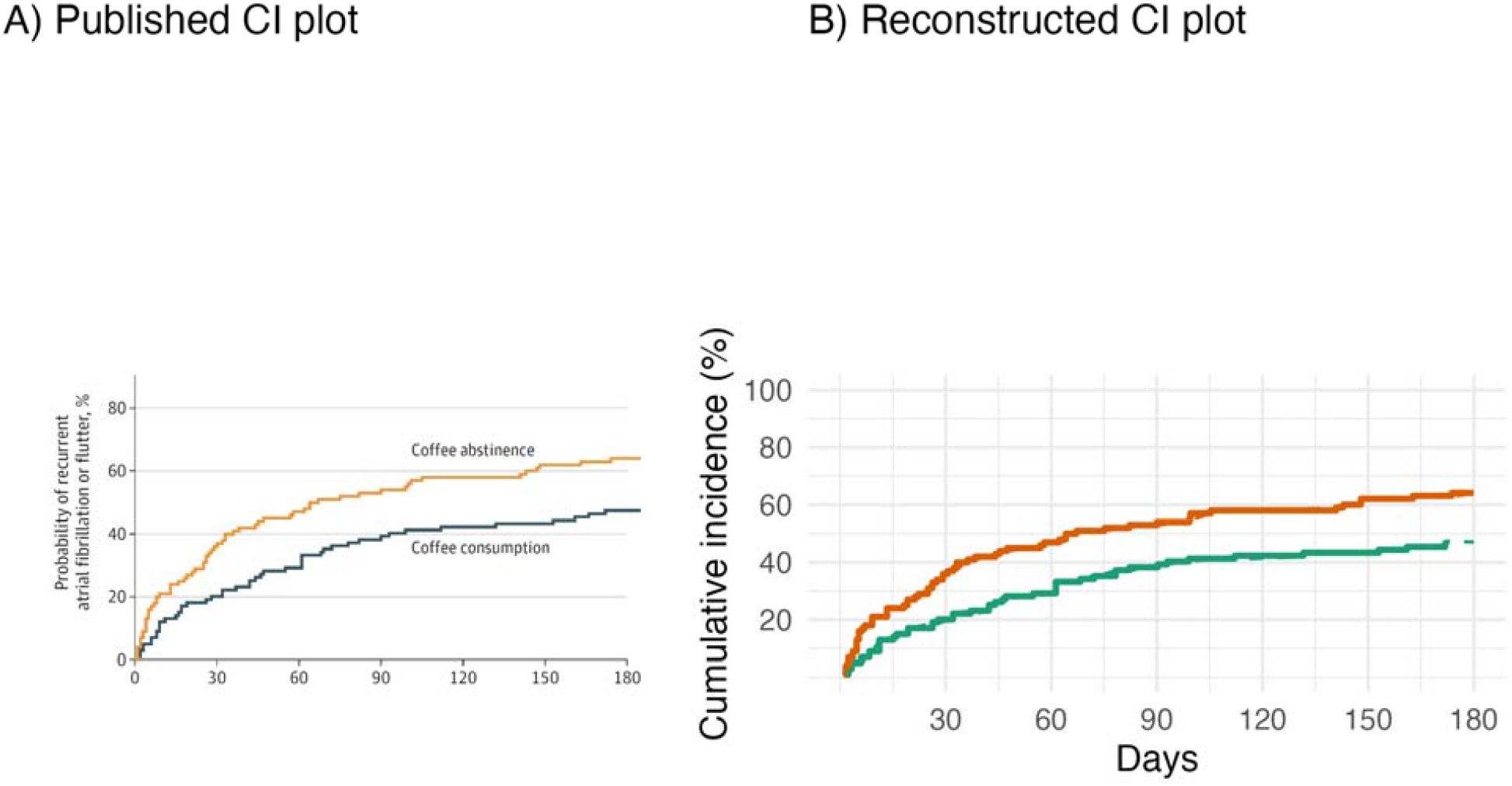
A) Cumulative incidence (CI) from JAMA DECAF publication B) Reconstructed cumulative incidence (CI) plot from digitalized JAMA DECAF curves

Once the priors and IPD are specified, we can proceed with the Bayesian survival regression modelling. As Cox regression uses the partial likelihood, which cancels out the baseline hazard when estimating the regression coefficients (i.e., log□HRs), in a Bayesian Cox survival model we only need a prior for the the treatment effect.

The Bayesian posterior hazard ratio is 0.74 (95% CrI 0.53–1.04), in favor of caffeinated coffee consumption being associated with a lower risk of AF recurrence compared to decaffeinated coffee. However compared to te published result, the strength of the association has been tempered by the prior belief that decaffeinated, and not caffeinated, coffee would be beneficial. Graphically this shift of the observed result by the prior belief is shown in Figure 4. Notice also the width of the posterior probability is narrowed compared to the prior distribution, underscoring that our new data has decreased our uncertainty about the treatment effect of coffee.

**Figure 4.**
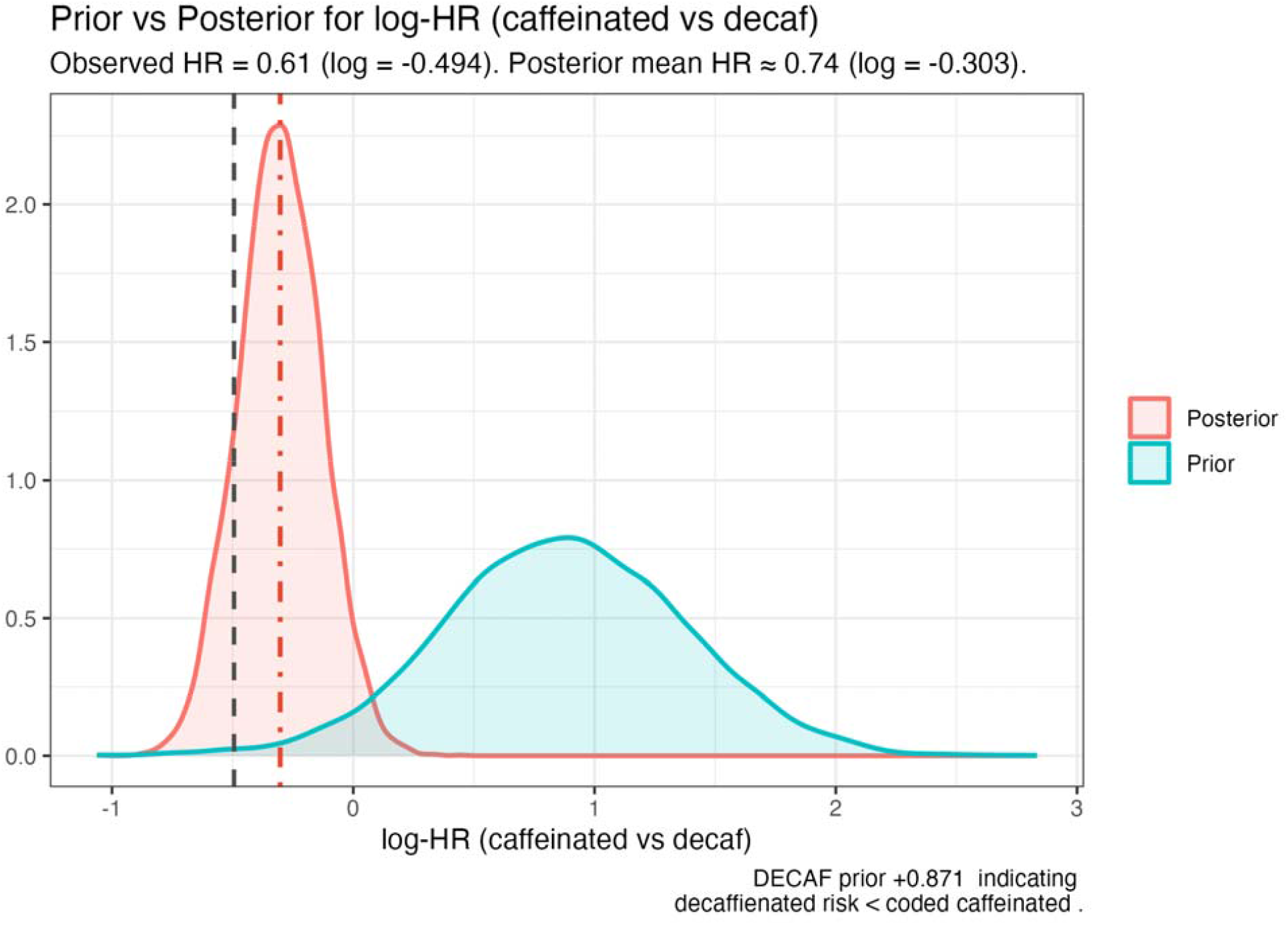
Prior and posterior probability density functions of log HR

In Figure 4, the dashed vertical line represents the observed frequentist log-HR of 0.61 (log = -0.494). The dot-dash vertical line represents the posterior mean log-HR of approximately 0.74 (log = -0.301). The prior distribution (teal) reflects a prior belief that decaffeinated coffee would be more beneficial than caffeinated coffee and therefore shifts the posterior distribution to the right of the observed value. The probability of a beneficial effect of caffeinated coffee (log HR <0 or HR < 1) is represented by the area under the curve (AUC) to the left of 0, which is 96%. However, the probability that caffeinated coffee has a clinically meaningful benefit, for example a HR < 0.9, falls to 88%. Increasing the threshold of what represents a clinically meaningful decrease, will lead to a corresponding reduction in the probability of a beneficial caffeine effect as determined by the DECAF(1) data.

### Bayesian risk difference

The DECAF(1) result can also be expressed in absolute terms as a risk difference, - 17% [95% CI; -.03, -.31, p = 0.01] and again a Bayesian approach may facilitate clinical interpretations. The risk difference prior, again based on the DECAF(1) power calculations, favored a decaffeinated benefit (89%) but did allocate a smaller probability (11%) for a possible caffeinated benefit. The Bayesian posterior risk difference is - 7.6% (95% CrI: - 19.5% to 4.4%), in favor of the caffeinated group. This posterior risk difference showed a shift towards the null compared to the frequentist result, due to the prior’s influence (Figure 5). While the final result showed a caffeinated coffee benefit, the strength of the evidence in favor of this benefit is seen as modest at best, and certainly less than implied by the original analysis. For example, the probability of a clinically meaningful benefit (e.g., say arbitrarily RD < -0.02 or number needed to treat = 50) is 82%.

**Figure 5.**
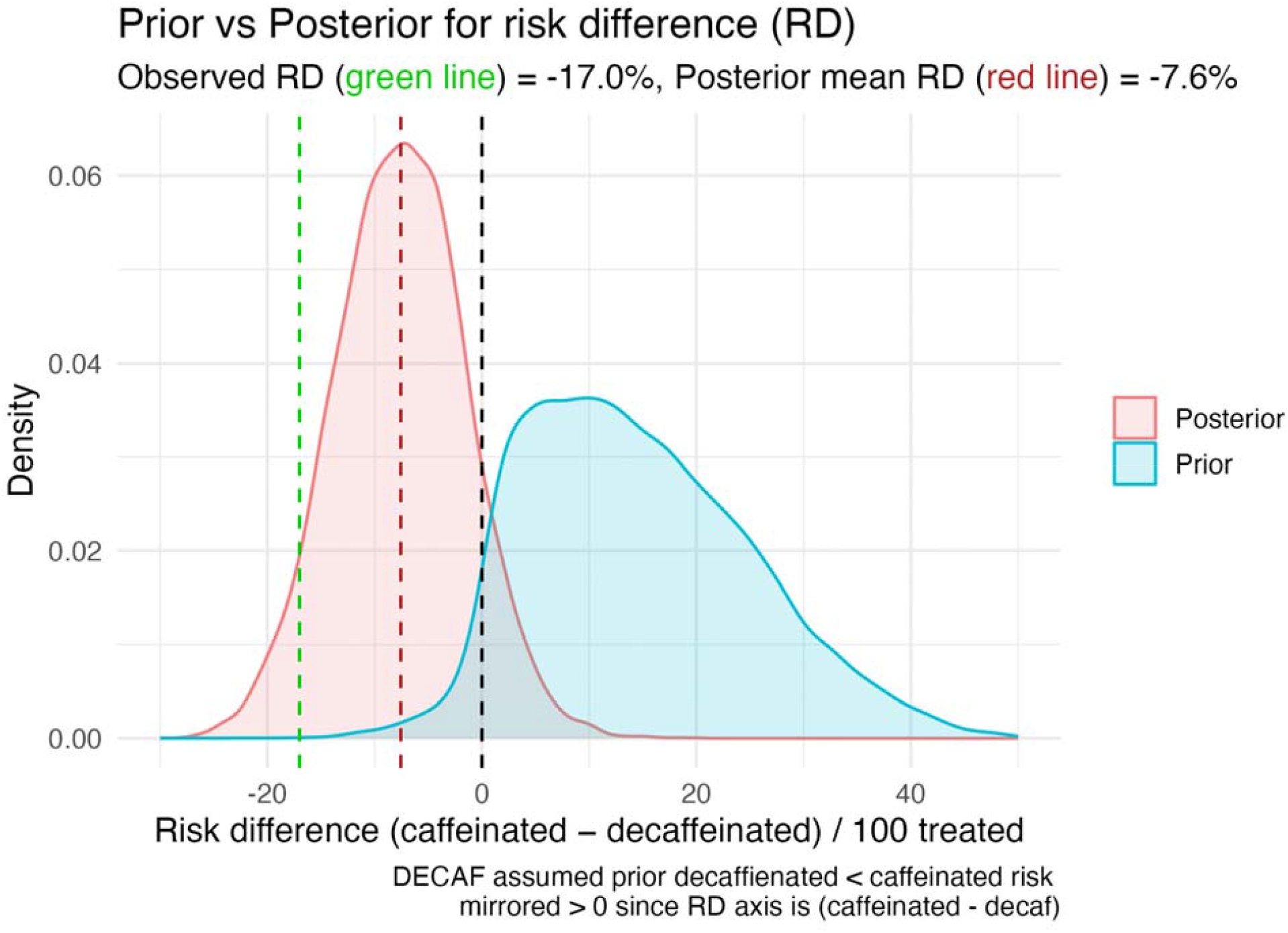
Prior and posterior probability density functions of risk difference

### Comparison of standard (frequentist) and Bayesian analyses

A comparison of the frequentist and Bayesian results are summarized in the Table. The main advantages of the Bayesian approach are the ability to incorporate prior beliefs and to provide direct probability estimates of benefit or harm, both overall and at clinically relevant thresholds. As shown, due to historical beliefs (context), the Bayesian reanalysis tempers the strength of evidence favoring caffeinated coffee compared to the frequentist interpretation. Moreover the Bayesian framework allows for a clearer distinction between statistical and clinical significance, which is crucial for informed decision-making.

### Are DECAF’s unexpected results a “one-off” phenomena?

Is DECAF(1) with its unexpected results an “one-off” case or do RCTs often report unexpected results that challenge prior beliefs. A small, convenience sample from three high impact journals (NEJM, JAMA, Lancet) found six superiority RCTs, published in 2024-25, with sample sizes ranging from 700 to 7000, whose results were in contradiction to prior beliefs as summarized in their pre-trial power calculations (see Supplemental File). All six trials based their final conclusions uniquely on their observed data, completely ignoring previous knowledge.

## Discussion

DECAF(1) reported a very strong, but surprising, association of decreased atrial fibrillation episodes in the caffeinated group (p = .01), but adjunctive frequentist and Bayesian analyses suggest this association is not conclusive. The small sample size and low statistical power to detect realistic effect sizes can lead to a M type exaggeration error. As the direction of improvement wasn’t specified, the potential for researcher’s degrees of freedom bias(15) also can’t be excluded. Most importantly, prior beliefs about the effect of caffeinated versus decaffeinated coffee were ignored.

Incorporating prior beliefs with a Bayesian reanalysis tempers the strength of evidence for the surprising result. For example, the probability of a clinically meaningful absolute risk difference of 2% is only approximately 80%. This distinction between statistical and clinical significance is crucial for informed decision-making and is sharpened within a Bayesian framework.

Equally surprising given the randomized study design and statistically significant result, was the authors’ interpretation that caffeinated coffee consumption was only “associated” and not casually related to less AF recurrence compared with abstinence from coffee and caffeinated products. Perhaps the authors like many clinicians preferred to follow their intuition or “gut instinct” and not encourage coffee consumption or maintenance as a preventive strategy for recurrent AF episodes. However, reliance on intuition over evidence is prone to cognitive errors(16) and hence our current infatuation with “dataism” - the belief that empirical data should guide our decisions as characterized by the pithy aphorism “In God we trust. All others have to bring data.”

Given this increasing influence of data-driven decision-making, it is essential that statistical interpretations be both rigorous and nuanced, whether frequentist or Bayesian in nature. From a frequentist standpoint, the performance of realistic sample size calculations is essential. DECAF’s(1) sample size was insufficient to detect realistic effect sizes, only ∼24% power to detect a 15% relative risk reduction, and hence any statistically significant result from such an underpowered study was likely to exaggerate the true effect — potentially by a factor of two.

DECAF’s small p-value informs us of the probability of observing this or more extreme data, if there was no true effect, and can be seen as unlikely. The inverse probability, the probability of an effect given the data, is arguably of greater interest and provides additional insights, most importantly the ability to include prior knowledge and to provide probability estimates of benefit or harm. Priors are often deemed the Achilles’ heel of these Bayesian analyses, but well justified, transparent and robust testing of different priors can mitigate subjectiveness concerns.

By incorporating prior beliefs and quantifying uncertainty in a way that frequentist methods cannot, the Bayesian framework adds further depth to the analysis. It enables clinicians to assess not only the most recently observed data but also how to contextualize it by incorporating existing knowledge and beliefs. This is particularly important in scenarios where prior evidence or expert opinion may conflict with new findings. Using priors derived from the DECAF authors’ own pre-trial assumptions(1), the Bayesian survival and risk difference models suggest that the strength of evidence favoring caffeinated coffee is more modest than the original frequentist interpretation has implied. As shown in the supplemental file, a conflict between RCT hypotheses and observed results is not rare and incorporating Bayesian methods may avoid fluctuating interpretations(17–19).

Some may dismiss this reanalysis as statistical alchemy or the “haze of Bayes”(20), but it offers a principled approach to contextualizing unexpected findings. For the DECAF(1) study, it tempers over-interpretation by including past beliefs, distinguishes between statistical and clinical significance, and provides the statistical justification for a replication study. This Bayesian reanalysis can help clinicians make more informed decisions by providing a clearer understanding of the evidence at hand. This is especially important given that over 120 media outlets reported the initial findings within a week of DECAF’s publication, potentially influencing clinical practice and public behavior based on an underpowered study with unexpected, and naively interpreted, results.

In conclusion, it is disconcerting that various DECAF(1) design issues were overlooked by authors, peer reviewers, journal editors, and national funding agencies. This reanalysis highlights the importance of rigorous statistical interpretation, whether frequentist or Bayesian, and underscores the need for careful consideration of prior beliefs and clinical significance in the interpretation of all trial results but particularly those with “unexpected” or “surprising” results. Future clinical trials should ensure adequate power, pre-specify analysis plans, and consider Bayesian approaches to provide a more comprehensive understanding of treatment effects.

**Tables.** Frequentist vs Bayesian comparison of DECAF results.

Frequentist vs Bayesian comparison of DECAF results
| Metric / Outcome | Frequentist (DECAF) | Bayesian Posterior |
| --- | --- | --- |
| Hazard Ratio (caffeinated vs abstinent) | HR = 0.61 (95% CI 0.42–0.89); $p \approx 0.01$ | HR ≈ 0.74 (95% CrI 0.53–1.04) |
| Risk Difference (caffeinated - abstinent) | RD ≈ -17.0% (95% CI -30.6% to -3.4%) | RD ≈ -7.6% (95% CrI -19.5% to +4.4%) |
| Pr(HR < 1) | — | ~96% |
| Pr(HR < 0.9) (clinical threshold example) | — | ~88% |
| Pr(RD < -0.02) (NNT ≤ 50 threshold example) | — | ~82% |

*Note:* Frequentist estimates are from the DECAF trial's published analysis; Bayesian posteriors use priors based on the original power assumptions and reconstructed IPD.

## Data Availability

All data produced are available online at https://github.com/brophyj/decaf.git

https://github.com/brophyj/decaf.git

## Supplementary Material

The Supplemental file that reviews recent RCTs with surprising results has been uploaded.

All code for the calculations and figures presented in this article are available online (https://github.com/brophyj/decaf.git).

