## supplemental file for "Understanding unexpected results from randomized clini□cal trials Does coffee reduce atrial fibrillation recurrences?"

Surprising Superiority RCTs (2024–2025)

James M Brophy

2026-01-03

### Supplemental Table

The DECAF trial(1), which produced a surprising result contradicting its original hypothesis, is not a “one-off” outlier. This table summarizes recent randomized controlled trials published in high-impact journals that were powered to detect specific intervention benefits but instead reported contradictory results(2–7).

Notably, authors’ conclusions consistently ignored prior expert opinions and earlier RCTs that informed their power calculations and hypothesized effects. Instead, they focused solely on observed results—a practice that risks erroneous conclusions and misguided clinical recommendations.

For example, the CLEAR trial(2) was powered to detect a 25% relative risk reduction with colchicine, assuming a 9% baseline event rate. Instead, it reported a neutral result (HR 0.99; 95% CI, 0.85–1.16; P = 0.93), leading authors to conclude colchicine offered no benefit after acute coronary syndrome. Yet, just three months later — without new evidence — the same investigators co-authored a meta-analysis claiming colchicine reduced major cardiovascular events by 12%(8). A Bayesian reanalysis(9) underscores how ignoring prior evidence and its heterogeneity fosters conflicting interpretations and potentially harmful clinical recommendations.

**Table 1 Trials (2024-2025) powered for benefit where observed results contradicted the hypothesis**

| Trial | Reference | Power calculations + hypothesized effect | Basis for Hypothesized Effect | Surprising result | Authors' conclusions |
| --- | --- | --- | --- | --- | --- |
| CLEAR SYNERGY (OASIS‑9): Colchicine vs placebo after MI (factorial RCT) | NEJM 2025 | Sample size: 3528 vs 3534 80% power to detect hypothesized 25% RRR (control ~9%) with colchicine | Previous RCTs and meta-analyses (COLCOT, LoDoCo2) and mechanistic rationale (CRP reduction) | Hazard ratio: 0.99 (95% CI, 0.85 to 1.16; P = 0.93) | Colchicine after MI did not reduce major adverse cardiovascular events |
| HEMOTION: Liberal vs restrictive transfusion in traumatic brain injury | NEJM 2024 | Sample size: 371 vs 371 80% power to detect hypothesized effect: −10% absolute difference with liberal policy | Expert opinion, prior observational/small RCT signals | Risk difference restrictive vs liberal: 5.4 percentage points (95% CI, −2.9 to 13.7) | Liberal transfusion strategy did not significantly reduce unfavorable outcomes |
| ENGAGES: EEG‑guided anesthesia vs usual care to prevent delirium after cardiac surgery | JAMA 2024 | Sample size: 567 vs 573 90% power to detect hypothesized effect: −8% absolute reduction with EEG-guided reduces delirium vs usual care | Prior RCTs and observational data linking EEG suppression to delirium | Risk difference: 0.05% (95% CI, −4.57% to 4.67%) | EEG-guided anesthesia did not reduce postoperative delirium |
| ARCADIA: Apixaban vs aspirin to prevent recurrent stroke in atrial cardiopathy without AF | JAMA 2024 | Sample size 507 vs 508 80% power to detect hypothesis apixaban 40% reduction in stroke vs aspirin | RCT evidence for anticoagulation effectiveness in AF, extrapolated to atrial cardiopathy | annualized risk difference in recurrent stroke 0% 95% CI −1.9 to 1.9 | Apixaban did not reduce recurrent stroke |
| ACHIEVE: Spironolactone vs placebo in maintenance dialysis (CV outcomes) | Lancet 2025 | Sample size spironolactone (n=1260) or placebo (n=1278, 90% powerr to detect a 25% reduction | Expert opinion | HR 0·92 (0·78–1·09); p=0·35 | Spironolactone did not reduce CV events |
| BedMed: Bedtime vs morning antihypertensive dosing to reduce CV events | JAMA 2025 | Sample size: 1677 vs 1680 80% power to detect hypothesis of 25% risk reduction : bedtime dosing reduces composite CV events vs morning dosing | Expert opinion, 2 previous RCTs | hazard ratio, 0.96; 95% CI, 0.77-1.19; P = .70) | Bedtime administration did not reduce CV events or death compared with morning dosing. |
| DECAF: Caffeinated coffee vs abstinence after cardioversion in AF/AFL | JAMA | Sample size: 100 vs 100 Hypothesized effect: not explicitly reported (see SAP at NCT05121519) | Expert caution and observational signals that caffeine may trigger AF; first RCT testing continuation vs abstinence | Neutral or harm presumed (coffee triggers AF) → benefit observed (↓ recurrence) | Continuing modest caffeinated coffee was associated with fewer AF/AFL recurrences over 6 months compared with abstinence, without excess adverse events. |
